# Automated Segmentation of Cerebral Arteries on Three-Dimensional Rotational Angiography Using nnUNet v2: Prospective Validation with Quantitative Metrics and Expert Qualitative Assessment

**DOI:** 10.64898/2026.05.20.26353640

**Authors:** Jeremy Hofmeister, Olivier Brina, Andrea Rosi, Gianmarco Bernava, Philippe Reymond, Michel Muster, Karl-Olof Lovblad, Paolo Machi

**Author notes:** **Corresponding author:** Dr Jeremy Hofmeister, Service de Neuroradiologie Diagnostique et Interventionnelle, Hôpitaux Universitaires de Genève, Geneva, Switzerland.

## Abstract

**Background:** Three-dimensional visualization and quantitative analysis of cerebral arteries on 3DRA are central to endovascular treatment planning, device selection, and cerebrovascular research. Manual segmentation is time-consuming and operator-dependent, yet no open-source deep learning model has been prospectively validated for this task on 3DRA.

**Methods:** A nnUNet v2 model was trained for binary cerebral artery segmentation on 400 consecutive 3DRA acquisitions from three angiographic systems, comparing four configurations across architectures and loss functions. The best-performing configurations were prospectively validated on 40 patients using a dual approach: quantitative metrics (DSC, clDice, HD95, ASD, Precision, Recall), and blinded expert qualitative evaluation by two interventional neuroradiologists assessing 12 arterial segments, a global quality score, and clinical usability across 40 test cases.

**Results:** The ensemble model achieved median DSC 0.917, clDice 0.932, and HD95 1.494 mm. Global quality scores were significantly lower for nnUNet v2 than for expert segmentations (median 4 vs 5, p<0.001), but nnUNet v2 segmentations were rated clinically usable in 88–90% of cases versus 95–98% for expert segmentations, without significant difference on the binary usability criterion. A consistent proximal-to-distal quality gradient was identified, with comparable scores at proximal arteries and the largest differences at distal arterial segments.

**Conclusion:** nnUNet v2 with topology-aware training provides clinically usable cerebral artery segmentations on 3DRA, prospectively validated through both quantitative metrics and structured expert qualitative assessment, and represents a reproducible open-source foundation for endovascular and research applications.

## Introduction

Neurovascular diseases, including intracranial aneurysms, arteriovenous malformations, dural arteriovenous fistulas, and ischemic stroke, represent major causes of morbidity and mortality worldwide.[1] Endovascular treatment has become the standard of care for an increasing proportion of these conditions, requiring precise knowledge of the neurovascular anatomy. Segmentation of cerebral arteries from medical imaging supports treatment planning, endovascular device selection and sizing, and computational fluid dynamics simulations. Beyond its immediate clinical role, reliable automated segmentation enables large-scale quantitative analysis of cerebrovascular morphology, with potential applications in the study of neurovascular pathologies and their relationship to treatment outcomes. Manual segmentation is time-consuming, operator-dependent, and poorly standardized, a barrier to both clinical use and large-scale research.[2,3] To date, no open-source, prospectively validated deep learning model exists for the automated segmentation of the complete cerebral arterial tree on three-dimensional rotational angiography (3DRA).

Three-dimensional rotational angiography (3DRA) is the reference 3D imaging modality for cerebrovascular intervention, often acquired during diagnostic cerebral angiography prior to endovascular treatment and offering superior spatial resolution and vascular contrast compared with magnetic resonance angiography and computed tomography angiography.[4,5] It is on these images that the interventional neuroradiologist plans the treatment and selects endovascular devices, making 3DRA the modality where automated segmentation would have the greatest direct clinical impact. However, the specific image characteristics of 3DRA prevent direct transfer of models trained on other modalities.

Deep learning has shown strong performance for cerebral vessel segmentation on other imaging modalities, with several open-source frameworks achieving high overlap and topological continuity metrics.[6] On 3DRA specifically, existing methods are either not open-source, not prospectively validated, or limited to specific pathologies.[4,7,8] Furthermore, most studies report performance exclusively through quantitative metrics, without evaluating clinical usability as perceived by the operators who perform endovascular procedures.

We aimed to develop and validate a state-of-the-art deep learning tool based on nnUNet v2 for automated segmentation of the complete cerebral arterial tree on 3DRA, trained on 400 acquisitions and prospectively validated on an independent cohort of 40 patients with anterior circulation intracranial aneurysms. Validation combined quantitative performance metrics with a structured expert qualitative assessment by two interventional neuroradiologists, evaluating segmentation quality at the arterial segment level and clinical usability for endovascular treatment planning and cerebrovascular anatomy analysis.

## Material & Methods

### Study design and patient cohorts

This study aimed to develop and validate a deep learning model based on nnUNet version 2 (v2) for the automated binary segmentation of cerebral arteries on three-dimensional rotational angiography (3DRA) data. The study comprised two distinct phases: a retrospective training phase and a prospective validation phase. The training cohort included 400 consecutive patients who underwent cerebral digital subtraction angiography with 3DRA acquisition at our institution between 2017 and 2022. Images were acquired on three angiographic systems: a Philips Azurion, a Philips Allura Clarity (Philips Healthcare, Best, The Netherlands), and a Siemens Artis Zee (Siemens Healthineers, Erlangen, Germany). Reconstructed 3DRA volumes had isotropic voxel sizes ranging from 0.12 to 0.40 mm and matrix sizes between 256^3^ and 512^3^ voxels. The prospective test cohort included 40 consecutive patients who underwent cerebral angiography with 3DRA acquisition for the initial workup of an anterior circulation intracranial aneurysm between January and April 2025. This cohort was entirely independent from the training dataset and was used exclusively for model performance evaluation. The study was approved by the local ethics committee (CCER 2017-00922).

### Reference standard and ground truth generation

All reference segmentations were performed manually using 3D Slicer (version 5.8.1; www.slicer.org). Segmentation was carried out through semi-automatic thresholding of the 3DRA volume followed by voxel-by-voxel manual correction by an expert. All intracranial arteries visible on the 3DRA volume were included in the segmentation, and a binary label was assigned to each volume (cerebral arteries = 1, background = 0). Three experts participated in ground truth generation: two interventional neuroradiologists and one interventional neuroradiology technician. To assess inter-expert reliability, the three experts independently segmented the same 40 cases, which were a subset of the training cohort. The Dice Similarity Coefficient (DSC) between raters ranged from 0.91 to 0.94, indicating good inter-expert agreement. Given this level of concordance, the segmentation produced by the most senior expert was retained as the reference label for these 40 cases. The remaining 360 cases were distributed among the three experts and segmented by a single expert per case, without label fusion or majority voting. For the test cohort, all 40 cases were segmented exclusively by the most senior expert to provide a consistent reference standard for quantitative performance evaluation.

### nnUNet v2 training

Model training was performed using nnUNet v2, an open-source self-configuring deep learning framework for biomedical image segmentation.[9,10] Preprocessing and network planning were performed using the nnUNetv2_plan_and_preprocess command, which automatically determined the optimal resampling, normalization, patch size, and batch size based on the dataset characteristics. The imaging modality was defined as angiography, resulting in Z-score normalization applied per volume. Four training configurations were compared. Configuration A used the standard 3d_fullres UNet architecture with the default Dice plus Cross-Entropy loss (DC+CE), serving as the baseline. Configurations B, C, and D used the Residual Encoder UNet Large preset (ResEncUNet L), planned with a GPU memory target of 32 GB. Configuration B used the same DC+CE loss as Configuration A. Configuration C added a soft centerline Dice loss (soft-clDice) to the DC+CE loss.[11] Configuration D replaced soft-clDice with the centerline boundary Dice loss (cbDice), which extends clDice by incorporating boundary-awareness.[12] All other training parameters were identical across configurations and followed nnUNet v2 defaults: stochastic gradient descent with Nesterov momentum of 0.99, polynomial learning rate decay, and the default on-the-fly data augmentation pipeline including rotations, elastic deformations, scaling, intensity augmentation, and mirroring. Each configuration was trained for 1000 epochs across five folds of cross-validation, with random case assignment to folds. The checkpoint with the highest exponential moving average pseudo-Dice on the validation set (checkpoint_best) was retained for each fold.

### Model selection

The best-performing configuration was identified using the nnUNetv2_find_best_configuration command with the --disable_ensembling option, which evaluated each configuration independently based on its mean cross-validation performance. The primary selection criterion was mean clDice across five validation folds, with mean DSC and mean HD95 used as secondary and tertiary criteria, respectively. If two configurations differed by less than 0.5% in mean clDice, a multi-configuration ensemble was tested using nnUNetv2_ensemble, which averages softmax outputs across folds before the final argmax. The final model consisted of the five-fold ensemble of the selected configuration, using checkpoint_best for inference in each fold.

### Quantitative performance evaluation

Quantitative model performance was evaluated on the prospective test cohort (n=40) by comparing the automated segmentation produced by the final model against the expert reference standard. Six metrics were computed globally on the full vascular mask, without segment-level analysis.[13] Overlap metrics included DSC, Precision, and Recall. Distance metrics included the 95th percentile Hausdorff Distance (HD95) and the Average Surface Distance (ASD). The centerline Dice coefficient (clDice) was computed as the primary topological metric using the official implementation.[11] All metrics are reported as median with interquartile range (IQR) and as mean with standard deviation (SD).

### Qualitative expert evaluation

The qualitative evaluation was performed on the test cohort by two experienced interventional neuroradiologists who perform endovascular treatments in routine clinical practice. Neither evaluator participated in ground truth generation. For each of the 40 patients, both the expert segmentation and the nnUNet segmentation were presented to each evaluator in a randomized order. Evaluators were blinded to the segmentation method and assessed the two segmentations independently, without knowledge of the other evaluator’s scores. Each segmentation was visualized in 3D Slicer v5.8.1 using a standardized protocol with identical orientation and volume rendering threshold. Twelve arterial segments were scored individually on a five-point Likert scale (1 = poor, 2 = insufficient, 3 = acceptable, 4 = good, 5 = excellent): four segments of the internal carotid artery (ICA: petrous, cavernous, ophthalmic, and communicating), four segments of the middle cerebral artery (MCA: M1 through M4), and four segments of the anterior cerebral artery (ACA: A1 through A4). A “not evaluable” (NE) option was available when a segment was absent, outside the field of view, or not identifiable. A global quality score was also assigned on the same five-point scale. Clinical usability was assessed using a four-level ordinal scale: A, no segmentation error, suitable for direct use in clinical practice and research; B, rare minor errors, acceptable for direct clinical use but requiring verification before use in research or quantitative vascular anatomy analysis; C, significant errors on some arterial segments, requiring manual correction before any use; D, unacceptable segmentation requiring major manual correction.

### Statistical analysis

Quantitative metrics are reported descriptively as median with IQR and mean with SD. No inferential testing was performed on quantitative metrics, as the expert segmentation constitutes the reference standard by definition. For the qualitative evaluation, the nnUNet and expert segmentations were compared using the Wilcoxon signed-rank test for each Likert item. The primary analysis included the global quality score and the clinical usability item. Secondary analyses compared territory-level scores, defined as the mean of individual segment scores within each territory (ICA, MCA, and ACA). Exploratory analyses reported scores for each of the 12 individual segments without inferential testing to avoid inflated type I error. Inter-rater agreement was assessed using the intraclass correlation coefficient (ICC, two-way mixed model, absolute agreement) for the global score, each territory score, and the clinical usability item, calculated on evaluable items only, excluding NE ratings. The proportion of segmentations rated as clinically usable without correction (categories A and B combined) was reported as a percentage for each method. A two-sided p-value below 0.05 was considered statistically significant. No correction for multiple comparisons was applied to secondary and exploratory analyses given their descriptive intent. Statistical analyses were performed using Python [3.14.1] and scipy.stats (Wilcoxon)[1.16.0] et pingouin (ICC)[0.6.1].

## Results

### Model selection and cross-validation performance

Four training configurations were evaluated by five-fold cross-validation on the training cohort (n=400). Configuration A, using the standard 3d_fullres UNet architecture with the default Dice plus Cross-Entropy loss, served as the baseline. Configurations B, C, and D used the ResEncUNet L architecture with increasing loss function complexity. Performance improved progressively across configurations on all metrics (Table 1). The mean clDice increased from 0.851 (Configuration A) to 0.923 (Configuration D), and the mean HD95 decreased from 3.211 mm to 1.891 mm over the same range. The difference in mean clDice between Configurations C and D was 0.4%, below the pre-specified 0.5% threshold, and both configurations were combined into a five-fold softmax ensemble. The ensemble achieved a mean clDice of 0.931 (SD 0.095), a mean DSC of 0.907 (SD 0.098), and a mean HD95 of 1.435 mm (SD 0.642 mm), outperforming each individual configuration on all metrics and was retained as the final model.

**Table 1.** Cross-validation performance of the four training configurations and the final ensemble model.

| Configuration | Architecture | Loss function | clDice |  | DSC |  | HD95 (mm) |  | ASD (mm) |  |
| --- | --- | --- | --- | --- | --- | --- | --- | --- | --- | --- |
|  |  |  | Mean | SD | Mean | SD | Mean | SD | Mean | SD |
| Config A | 3d_fullres UNet | DC + CE | 0.851 | 0.102 | 0.817 | 0.103 | 3.211 | 0.988 | 0.418 | 0.121 |
| Config B | ResEncUNet L | DC + CE | 0.887 | 0.097 | 0.836 | 0.097 | 2.871 | 0.870 | 0.388 | 0.107 |
| **Config C** | **ResEncUNet L** | **DC + CE + soft-clDice** | **0.919** | **0.107** | **0.896** | **0.109** | **1.997** | **0.612** | **0.295** | **0.092** |
| **Config D** | **ResEncUNet L** | **DC + CE + cbDice** | **0.923** | **0.095** | **0.897** | **0.097** | **1.891** | **0.678** | **0.242** | **0.096** |
| Ensemble (C+D) | Configs C + D | 5-fold softmax avg | 0.931 | 0.095 | 0.907 | 0.098 | 1.435 | 0.642 | 0.226 | 0.093 |

Values reported as mean ± SD across 5 validation folds. Bold rows = configurations selected for ensemble testing (clDice difference 0.4%, below the pre-specified 0.5% threshold).
*Abbreviations:* DSC = Dice Similarity Coefficient; HD95 = Hausdorff Distance 95th percentile; ASD = Average Surface Distance; clDice = centerline Dice; DC = Dice loss; CE = Cross-Entropy loss.

### Quantitative performance on the test cohort

The final ensemble model was evaluated on the prospective test cohort of 40 patients with anterior circulation intracranial aneurysms. Quantitative performance metrics are reported in Table 2 and illustrated in Figure 1. The model achieved a median DSC of 0.917 (IQR 0.878–0.955), a median clDice of 0.932 (IQR 0.894–0.959), and a median HD95 of 1.494 mm (IQR 1.178–2.237 mm). Median Precision and Recall were 0.904 (IQR 0.867–0.951) and 0.918 (IQR 0.882–0.962), respectively, indicating a slight tendency toward under-segmentation over over-segmentation. The median ASD was 0.221 mm (IQR 0.177–0.324 mm). The HD95 distribution showed right-skewed asymmetry, with a maximum value of 5.171 mm driven by a small number of cases with segmentation errors on distal arterial segments.

**Table 2.** Quantitative segmentation performance of the final nnUNet v2 ensemble model on the prospective test cohort.

| Metric | Median | Q1 | Q3 | Mean | SD | Min | Max |
| --- | --- | --- | --- | --- | --- | --- | --- |
| **Overlap metrics** |  |  |  |  |  |  |  |
| DSC | 0.917 | 0.878 | 0.955 | 0.901 | 0.071 | 0.737 | 0.987 |
| Precision | 0.904 | 0.867 | 0.951 | 0.896 | 0.075 | 0.697 | 0.970 |
| Recall | 0.918 | 0.882 | 0.962 | 0.909 | 0.081 | 0.748 | 0.986 |
| **Distance metrics** |  |  |  |  |  |  |  |
| HD95 (mm) | 1.494 | 1.178 | 2.237 | 1.528 | 0.559 | 0.487 | 5.171 |
| ASD (mm) | 0.221 | 0.177 | 0.324 | 0.237 | 0.080 | 0.104 | 0.612 |
| **Topology metric** |  |  |  |  |  |  |  |
| clDice | 0.932 | 0.894 | 0.959 | 0.921 | 0.062 | 0.791 | 0.984 |

*Abbreviations:* DSC = Dice Similarity Coefficient; HD95 = Hausdorff Distance 95th percentile; ASD = Average Surface Distance; clDice = centerline Dice. Q1 = first quartile; Q3 = third quartile; SD = standard deviation.

**Figure 1:**
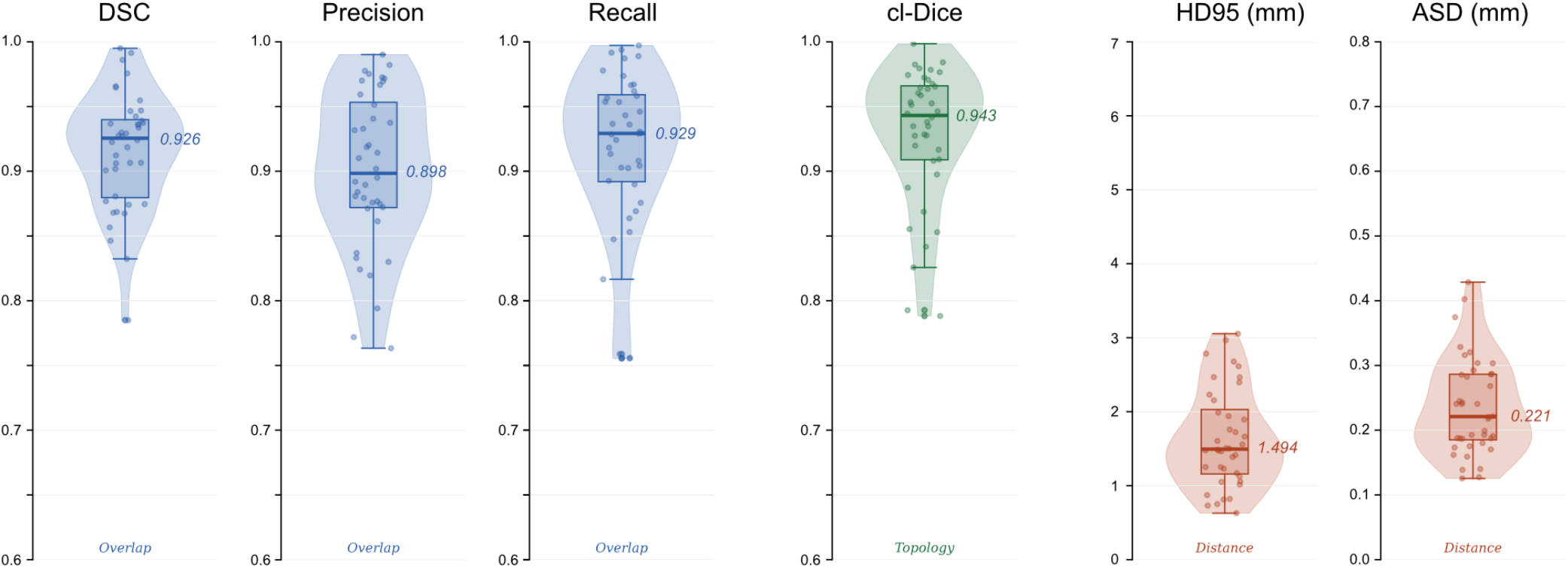
Distribution of quantitative segmentation metrics for the nnUNet v2 model on the prospective test cohort. Legend: Violin plots show the full distribution of values across 40 patients of the test cohort. Box plots display the median, interquartile range, and 1.5*IQR whiskers. Individual data points are overlaid with jitter. The median value is annotated to the right of each box. Metrics are grouped by type: overlap metrics (DSC, Precision, Recall; blue), topology metric (clDice; green) and distance metrics (HD95, ASD; red). Abbreviations: DSC = Dice Similarity Coefficient; HD95 = Hausdorff Distance at the 95th percentile; ASD = Average Surface Distance; clDice = centerline Dice; IQR = interquartile range.

### Qualitative expert evaluation Primary analysis

The global quality score and clinical usability item were assessed by two experienced interventional neuroradiologists for both the expert and nnUNet v2 segmentations across 40 patients. The global quality score was significantly higher for the expert segmentation than for the nnUNet v2 segmentation for both raters (Rater 1: median 5 [IQR 4–5] vs 4 [IQR 4–5], p<0.001; Rater 2: median 5 [IQR 4–5] vs 4 [IQR 4–5], p<0.001). Regarding clinical usability, the expert segmentation was rated as directly usable (category A or B) in 98% of cases by Rater 1 and 95% by Rater 2. The nnUNet v2 segmentation was rated as directly usable in 90% of cases by Rater 1 and 88% by Rater 2. The difference in clinical usability reached statistical significance for Rater 1 (p=0.011) but not for Rater 2 (p=0.134). Inter-rater agreement on the global quality score was good for the expert segmentation (ICC 0.777) and moderate for the nnUNet v2 segmentation (ICC 0.611). Inter-rater agreement on clinical usability was good for both methods (ICC 0.830 and 0.845, respectively).

### Secondary analysis

Territory-level scores are reported in Table 3 and illustrated in Figures 2 and 3. The nnUNet v2 segmentation received significantly lower scores than the expert segmentation across all three vascular territories for both raters (all p<0.001). For the ICA, the median territory score was 5 for the expert and 4.8 (Rater 1) or 4.5 (Rater 2) for nnUNet v2. Differences were larger for the MCA and ACA, where the expert median territory score was 4.5 for both raters, compared with 3.5 for nnUNet v2.

**Table 3.** Qualitative evaluation: Likert scores and clinical usability for expert and nnUNet v2 segmentations on the prospective test cohort.

|  | Expert segmentation |  | nnUNet v2 segmentation |  | p-value<sup>†</sup> (Expert vs nnUNet) |  | ICC* (Rater 1 vs Rater 2) |  |
| --- | --- | --- | --- | --- | --- | --- | --- | --- |
|  | Rater 1<br>Median<br>[Q1–Q3] | Rater 2<br>Median<br>[Q1–Q3] | Rater 1<br>Median<br>[Q1–Q3] | Rater 2<br>Median<br>[Q1–Q3] | Rater 1 | Rater 2 | Expert | nnUNet v2 |
| **PRIMARY ANALYSIS** |  |  |  |  |  |  |  |  |
| Global quality score | 5 [4–5] | 5 [4–5] | 4 [4–5] | 4 [4–5] | p<0.001 | p<0.001 | 0.777 | 0.611 |
| Clinical usability | A: 78%<br>B: 20%<br>C: 2%<br>D: 0% | A: 70%<br>B: 25%<br>C: 5%<br>D: 0% | A: 65%<br>B: 25%<br>C: 10%<br>D: 0% | A: 62%<br>B: 25%<br>C: 12%<br>D: 0% | p=0.011 | ns | 0.830 | 0.845 |
| **SECONDARY ANALYSIS - Vascular territories** |  |  |  |  |  |  |  |  |
| ICA (all segment) | 5 | 5 | 4.8 | 4.5 | p<0.001 | p<0.001 | 0.804 | 0.778 |
| MCA (all segment) | 4.5 | 4.5 | 3.5 | 3.5 | p<0.001 | p<0.001 | 0.720 | 0.765 |
| ACA (all segment) | 4.5 | 4.5 | 3.5 | 3.5 | p<0.001 | p<0.001 | 0.843 | 0.859 |
| **EXPLORATORY ANALYSIS - individual arterial segments** |  |  |  |  |  |  |  |  |
| ICA - Petrous | 5 [5–5] | 5 [5–5] | 5 [5–5] | 5 [5–5] | ns | ns | 0.649 | 0.310 |
| ICA - Cavernous | 5 [5–5] | 5 [4–5] | 5 [4–5] | 5 [4–5] | p=0.003 | ns | 0.459 | 0.603 |
| ICA - Ophthalmic | 5 [4–5] | 5 [4–5] | 4.5 [4–5] | 4 [4–5] | p=0.020 | p=0.003 | 0.943 | 0.712 |
| ICA - Communicating | 5 [5–5] | 5 [4.5–5] | 4 [4–5] | 4 [4–4] | p<0.001 | p<0.001 | 0.797 | 0.832 |
| MCA - M1 | 5 [5–5] | 5 [5–5] | 5 [5–5] | 5 [4.5–5] | ns | ns | 0.614 | 0.769 |
| MCA - M2 | 5 [4–5] | 5 [4–5] | 4 [4–5] | 4 [4–5] | p<0.001 | p=0.002 | 0.731 | 0.686 |
| MCA - M3 | 4 [4–5] | 4 [4–5] | 3 [3–4] | 3 [3–4] | p<0.001 | p<0.001 | 0.668 | 0.761 |
| MCA - M4 | 4 [3.5–4] | 3.5 [3–4] | 2 [2–3] | 2 [2–3] | p<0.001 | p<0.001 | 0.589 | 0.763 |
| ACA - A1 | 5 [5–5] | 5 [5–5] | 5 [4.5–5] | 5 [4.5–5] | p=0.008 | ns | 0.552 | 0.944 |
| ACA - A2 | 5 [5–5] | 5 [4.5–5] | 4 [3.5–4] | 4 [3.5–4] | p<0.001 | p<0.001 | 0.897 | 0.903 |
| ACA - A3 | 4 [4–5] | 4 [4–5] | 3 [2–3] | 3 [2–3] | p<0.001 | p<0.001 | 0.822 | 0.606 |
| ACA - A4 | 4 [3–5] | 4 [3–4] | 2 [2–3] | 2 [2–3] | p<0.001 | p<0.001 | 0.691 | 0.740 |

Values reported as Median [Q1–Q3] for individual segment Likert items (scale 1–5). Q1 and Q3 computed using the midpoint interpolation method. Vascular territory score = median of the per-segment medians (4 segments per territory); no IQR reported as this is a summary of summary statistics. Wilcoxon p-values for territory analyses computed on per-patient composite scores (mean of 4 segment scores per patient). Utilisability reported as percentage per category (A = no error, direct use; B = rare minor errors, direct clinical use; C = significant errors, correction required; D = unacceptable). † Wilcoxon signed-rank test (paired), nnUNet v2 vs. expert, per rater.
*Abbreviations:* ICC = intraclass correlation coefficient (two-way mixed model, absolute agreement), Rater 1 vs Rater 2, computed separately for expert and nnUNet v2 segmentations. ICA = internal carotid artery; MCA = middle cerebral artery; ACA = anterior cerebral artery.

**Figure 2:**
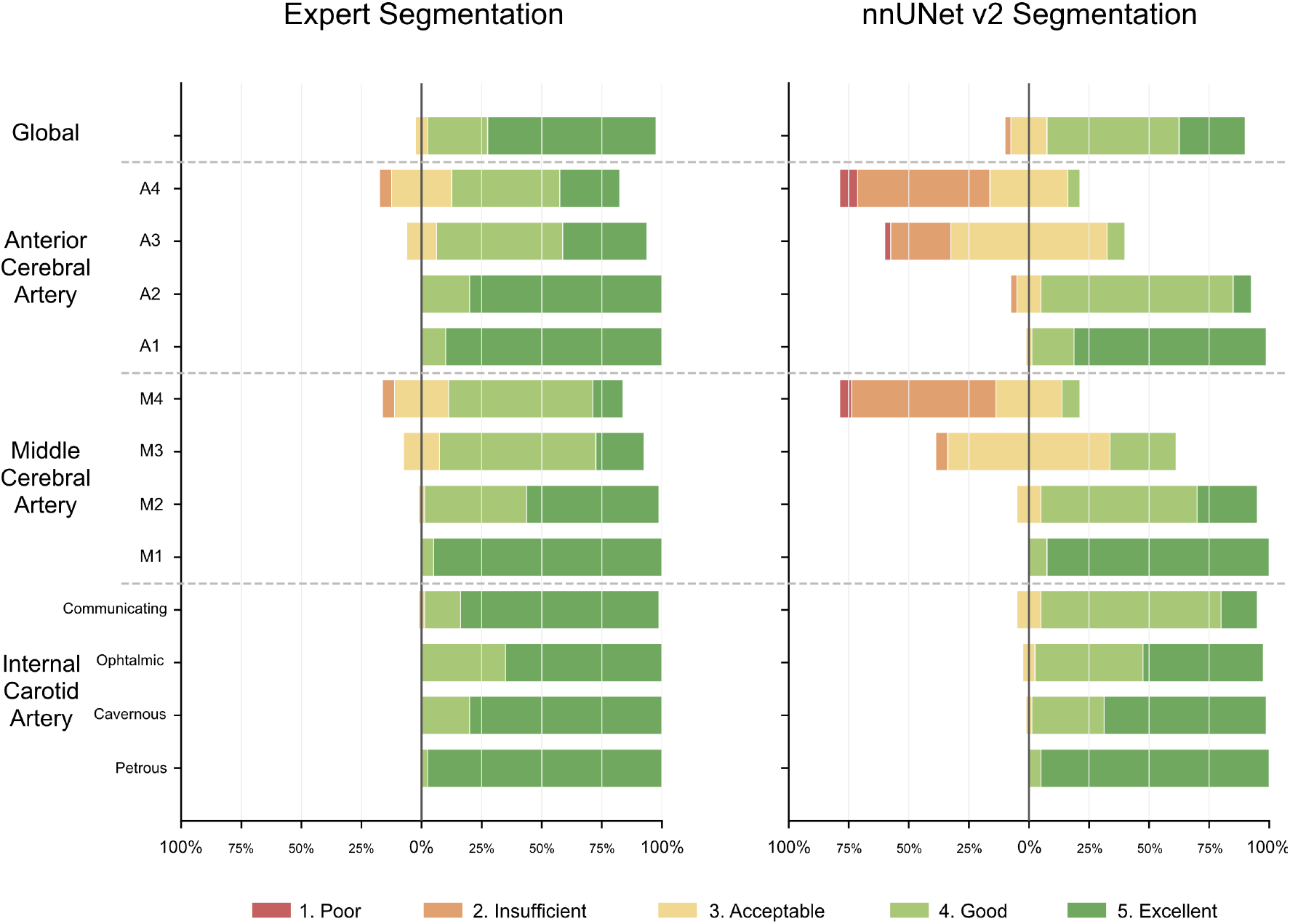
Qualitative Likert scores for expert and nnUNet v2 segmentations across 12 arterial segments and the global quality score. Legend: Each bar represents the distribution of scores (1–5) for a given segment and method, averaged across the 2 raters. Bars extending to the left of the central axis represent poor or insufficient scores (1–2); bars extending to the right represent good or excellent scores (4–5); the central bar represents acceptable scores (3).

**Figure 3:**
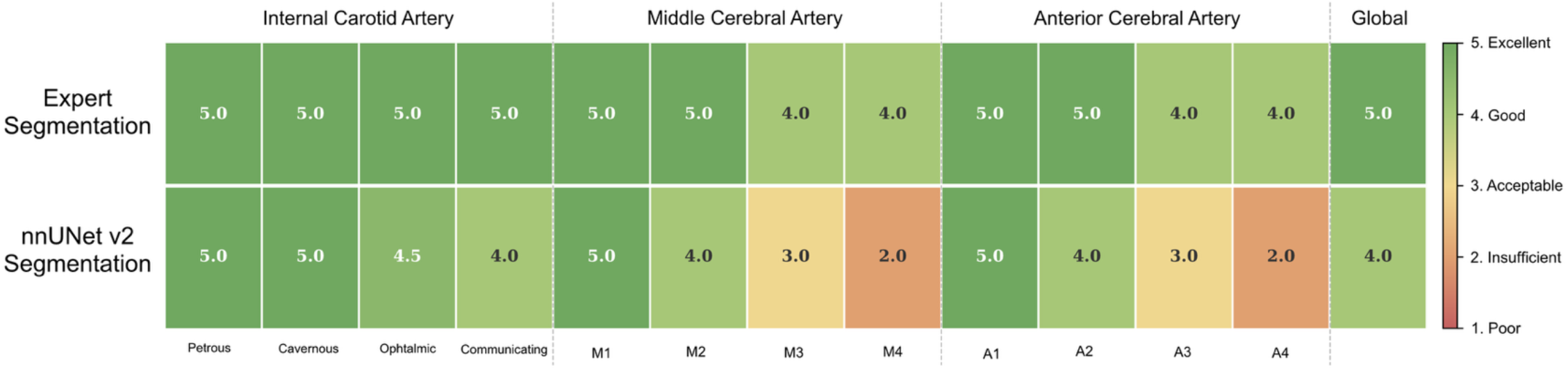
Likert scores (scale 1–5) for expert and nnUNet v2 segmentations across 12 arterial segments and the global quality score. Legend: Cell color reflects the median score on a 5-point Likert scale, ranging from poor (score 1, red) to excellent (score 5, green), averaged across the 2 raters. Numerical values within each cell indicate the median score (averaged across 2 raters).

### Exploratory analysis

Segment-level results are reported in Table 3. No significant difference was found between the expert and nnUNet v2 segmentations for the ICA petrous segment (both raters, p>0.05) or the MCA M1 segment (both raters, p>0.05). Significant differences were observed for the ICA communicating segment (both raters, p<0.001), all MCA segments from M2 onward, and all ACA segments from A2 onward. The largest score differences were found for the distal segments: median nnUNet v2 scores of 2 [IQR 2– 3] were observed for MCA M4 (both raters) and ACA A4 (both raters), compared with expert median scores of 4 [IQR 3.5–4] and 4 [IQR 3–5], respectively. A consistent proximal-to-distal gradient in segmentation quality was observed for nnUNet v2, while expert segmentation scores remained more uniform across segments (Figures 2 and 3). The main error patterns identified by the raters were incomplete segmentation of distal MCA and ACA branches (M3–M4 and A3–A4) and segmentation artifacts at the level of the ICA siphon in the presence of calcified atherosclerotic plaques, which occasionally resulted in false gaps or missing vessel segments in the cavernous and communicating portions. These errors were infrequent at the ICA level but contributed to the lower scores observed on the distal segments. Inter-rater agreement was good to excellent for most segments (ICC range 0.459–0.944), with the lowest values observed for the ICA petrous and cavernous segments, where a ceiling effect was present in both methods.

## Discussion

The primary aim of this study was to develop and prospectively validate a nnUNet v2-based model for automated binary segmentation of cerebral arteries on 3DRA, using both quantitative metrics and expert qualitative assessment. The final model, a five-fold ensemble combining the two best-performing configurations, achieved a median DSC of 0.917, a median clDice of 0.932, and a median HD95 of 1.494 mm on the prospective test cohort. In the qualitative evaluation, nnUNet v2 segmentations were rated as clinically usable without correction in 88–90% of cases, compared with 95– 98% for expert segmentations, with no statistically significant difference on the binary usability criterion.

### Comparison with prior work on 3DRA

Several groups have previously addressed vessel and aneurysm segmentation in 3DRA using deep learning. Lin et al. proposed a multi-class CNN for simultaneous vessel and aneurysm segmentation, achieving a vessel DSC of 0.91 and an average surface-to-surface error of 0.25 mm on a multi-center internal dataset.[7] Our model, trained on a binary segmentation task, which is inherently more tractable than multi-class segmentation, achieved comparable vessel DSC, while being based on an open-source, self-configuring framework that requires no deep learning expertise to deploy. Garcia et al. introduced a 3DUNet-based approach for vessel segmentation in 3DRA of brain arteriovenous malformations, reporting promising coverage of relevant structures despite a very limited training set of five annotated volumes.[14] Nishi et al. trained a model largely based on nnUNet v1 for cerebral aneurysm segmentation and morphological analysis on 3DRA, reporting a mean DSC of 0.87 for aneurysm segmentation.[4] While their approach extends beyond vessel segmentation to automated morphological parameter extraction, the vessel segmentation component was not the primary endpoint, and the model was not validated prospectively or evaluated by clinical raters. Our study addresses both gaps by providing a prospectively validated open-source model with structured expert qualitative assessment.

### Comparison with prior work on MRA-TOF and CTA

A recent study trained a nnUNet on MRA-TOF images of 62 patients and reported a median DSC of 0.86 across all segmented classes, including parent vessels and intracranial aneurysms, with a median clDice of 0.91 and a median HD95 of 2.9 mm.[15] Our model, trained on 3DRA, a modality with superior spatial resolution and vascular contrast, achieved higher DSC and clDice, and lower HD95, consistent with the expectation that higher image quality translates into improved segmentation performance. Orouskhani et al. similarly used nnUNet with various loss function combinations on TOF-MRA for intracranial aneurysm segmentation, demonstrating that hybrid loss strategies improve performance over the default Dice plus Cross-Entropy baseline.[16] Our findings corroborate this: the progressive improvement from Configuration A to the final ensemble, with topological loss functions contributing to the largest performance gains on clDice and HD95, confirms the value of topology-aware training for vascular tubular structures.

### Clinical and scientific utility

Automated segmentation of cerebral arteries on 3DRA has direct implications for clinical practice and research. In the endovascular context, accurate three-dimensional vascular models derived from 3DRA are essential for treatment planning, selection of therapeutic devices, and measurement of morphological parameters required for device sizing, including parent artery diameter, vessel tortuosity, and landing zone geometry.[17] Nishi et al. demonstrated that automated morphological analysis of cerebral aneurysms based on 3DRA data can provide clinically relevant parameters, including neck diameter and aneurysm height, with errors within acceptable clinical tolerances.[4] A reliable vessel segmentation model is a prerequisite for such downstream applications. Beyond the procedural setting, automated segmentation opens the way to quantitative analysis of cerebrovascular anatomy at population scale.

CaravelMetrics, a recent computational framework applied to 570 MRA datasets, demonstrated that automated graph-based morphometric analysis of cerebral vessels captures age- and sex-related variations in vascular complexity, providing a scalable approach for normative modeling and population-level studies of vascular health.[18] Comparable approaches have identified significantly increased tortuosity and fractality with age and in patients with acute ischemic stroke and Alzheimer’s disease, compared with healthy controls.[19] The superior spatial resolution of 3DRA, which captures distal vessel branches invisible on MRA or CTA, makes it particularly well suited for such morphometric research in patients already undergoing angiography for clinical indications. Houry et al. further demonstrated that vessel segmentation from 3DRA can generate planar vascular maps that preserve geometric properties relevant to catheter navigation, with potential direct integration into the endovascular workflow.[20]

### Limitations and perspectives

Several limitations of this study should be acknowledged. First, both the training and test cohorts were acquired at a single institution, which may limit the generalizability of the model to other angiographic systems or acquisition protocols. Second, the prospective test cohort was restricted to anterior circulation aneurysms, and performance on posterior circulation or other neurovascular pathologies remains to be evaluated. Third, the qualitative evaluation identified two main error patterns: incomplete delineation of distal MCA and ACA branches, and segmentation artifacts at the level of the ICA siphon in the presence of calcified atherosclerotic plaques. The former reflects a general limitation of binary vessel segmentation models on fine tubular structures at the boundary of image resolution, while the latter results from the altered intraluminal signal produced by calcified plaques, which disrupts the intensity contrast exploited by the thresholding-based ground truth and the model alike. Topological loss functions partially mitigated connectivity errors on distal branches, as reflected by the improved clDice scores, but did not fully resolve this challenge. Fourth, the current binary segmentation does not distinguish individual arterial segments, which limits direct segment-level quantitative analysis. Future work will focus on multi-class segmentation to enable per-segment morphometric measurements, external validation on multi-center datasets, and integration of vessel wall imaging to improve robustness in the presence of atherosclerotic disease.

## Conclusion

We trained and prospectively validated a nnUNet v2-based model for automated segmentation of cerebral arteries on 3DRA and found that a topology-aware ensemble combining centerline-based loss functions outperformed the standard baseline across all metrics. Expert qualitative evaluation confirmed that the model produced clinically usable segmentations in 88–90% of cases. This open-source, self-configuring approach provides a reproducible and prospectively validated baseline for automated cerebrovascular segmentation on 3DRA, with direct applications in endovascular treatment planning and quantitative vascular anatomy research.

## Data Availability

All data produced in the present study are available upon reasonable request to the authors.

## Notes

### Competing Interest Statement

The authors have declared no competing interest.

### Funding Statement

This study did not receive any funding.

### Author Declarations

The project was approved by the "Commission Cantonale d'Ethique de la Recherche sur l’être humain (CCER)" from the Geneva State, in Switzerland. It has the approval number: CCER 2017-00922.

